# KCL TEST: an open-source inspired asymptomatic SARS-CoV-2 surveillance programme in an academic institution

**DOI:** 10.1101/2023.07.25.23293154

**Authors:** J. Reis de Andrade, E. Scourfield, S. Peswani-Sajnani, K. Poulton, T. Ap Rees, P. Khooshemehri, G. Doherty, S. Ong, I. Ivan, N. Goudarzi, I. Gardiner, E. Caine, T. Maguire, D. Leightley, L. Torrico, A. Gasulla, A. Menendez-Vazquez, Ana Maria Ortega Prieto, Suzanne Pickering, Jose Jimenez Guardeño, Rahul Batra, A.V.F. Tan, A. Griffin, S. Papaioannou, C. Trouillet, H. Mischo, V. Giralt, S. Wilson, M. Kirk, Stuart Neil, Rui Pedro Ribeiro Galao, J. Martindale, C. Curtis, M. Zuckerman, R. Razavi, M.H. Malim, R.T. Martinez-Nunez

## Abstract

Testing was paramount in the management of the COVID-19 pandemic. Our university established KCL TEST: a SARS-CoV-2 asymptomatic testing programme that enabled sensitive and accessible PCR testing of SARS-CoV-2 RNA in saliva. We performed 158,277 PCRs for our staff, students, and their household contacts, free of charge. Our average turnaround time was 16h and 37 mins from user registration to result delivery. KCL TEST combined open-source automation and in house non-commercial reagents, which allows for rapid deployment. Here we provide our blueprint, recently recommended for ISO15189 accreditation, and results to enable the rapid launch of diagnostic laboratories where and when needed, particularly in low-resource settings. Our data span over 18 months and parallels that of the UK Office for National Statistics, with a lower positive rate and virtually no delta wave. Our observations strongly support regular asymptomatic community testing to decrease outbreaks and provide safe working spaces. Universities can therefore provide agile, resilient, and accurate testing that reflects the infection rate and trend of the general population. We call for the integration of academic institutions in pandemic preparedness, with capabilities to rapidly deploy highly skilled staff, as well as develop, test and accommodate efficient low-cost pipelines.

## Introduction

The rapid establishment of diagnostic facilities was essential to minimise the spread of SARS-CoV-2 in the community since the start of the COVID-19 pandemic. Diagnostic laboratories worldwide, mainly based at hospitals and clinics, were rapidly overwhelmed by the demand, in addition to the lack of reagents and supplies that we and many others reported ^1 2^. Mass testing programmes in very large laboratories (‘Lighthouse Labs’ in the UK) were in large part set up by volunteers from universities^3^. However, contact tracing of cases and management of such large facilities was complex, with sample turnaround times being days from sampling to result. Testing was mostly restricted to symptomatic individuals, while clear asymptomatic transmission happened in the community.

Academic institutions became hubs where scientists changed their day-to-day research and/or teaching jobs and contributed to testing in multiple countries worldwide ^4-9^. Our institution was no exception and we developed novel protocols based on reagents used in molecular research labs ^2^, repurposed research spaces into diagnostic facilities ^10^ and with many others contributed with economic, accessible and sensitive methods to allow testing in resource-limited settings ^11-15^. Universities have, proportionally, a large relatively young population (students), which had less risk of developing acute or severe COVID-19 as compared with older adults. However, Universities also faced challenges to manage outbreaks considering the living space in halls or residences, small offices and laboratories. Transmission can therefore be high ^16^, of particular relevance in shared spaces with hospitals. Even 18 months after the pandemic onset, testing at universities was proposed as advantageous ^17 18^. Additionally, academic centres also pose an opportunity to engage with a community that is familiar with research and can be more easily engaged in pilot testing programmes ^19^. We have a flexible workforce, space availability linked with research, as well as experience in service provision and management.

We set out to ensure a safe return to campus by providing SARS-CoV-2 RNA testing, with four main premises: high sensitivity and specificity, ease of use, rapid turnover of results and minimal costings. KCL TEST processed nearly 160,000 samples between December 2020 and July 2022. We tested saliva for SARS-CoV-2 RNA using PCR and mainly open-source automation and non-commercial molecular protocols, with an average turnaround time of 8 h and a limit of detection of 100 copies / mL. Between 10% and 30% of campus footfall was tested daily for 18 months. Our data were fed into the NHS Test and Trace programme, contributing to national testing efforts. KCL TEST has been recommended by the United Kingdom Accreditation Service (UKAS) for ISO15189 accreditation.

Our system allows for up- and down-scaling, making it flexible and deployable in multiple settings and for multiple purposes. We provide our blueprint and protocols, experience in accreditation and data, as a guide to help others navigate the agile set-up of new laboratories. We believe our data also highlight the value of asymptomatic testing in the community, using a simpler sample to test. We propose that Governments should engage with academic centres as soon as the need for widespread testing is apparent, to enable rapid development and testing of pipelines and alleviate overstretched healthcare systems.

## Results

### KCL TEST overview

KCL TEST was comprised of multiple teams, covering operations, laboratory and management (Figure 1). We opted for PCR methodology due to high sensitivity and specificity, and saliva as a sample for its ease of collection. PCR analyses of saliva were considered as accurate and sensitive for SARS-CoV-2 detection by September 2020 ^20^, although it posed issues for automation in many laboratories. Saliva is more viscous than Universal Transport Medium (UTM) or Viral Transport Medium (VTM) used with swabs which can increase the chances of cross-contamination when pipetting by robots. By comparison, swab samples collected into UTM or VTM pose a risk during transportation and require inactivation of possible virus to protect laboratory personnel prior to commencing analysis. Sample collection directly into inactivation medium offered an alternative to UTM/VTM but required validations for sensitivity and specificity. We set out to overcome these barriers and provide an end-to-end validated system for SARS-CoV-2 testing.

**Figure 1.**
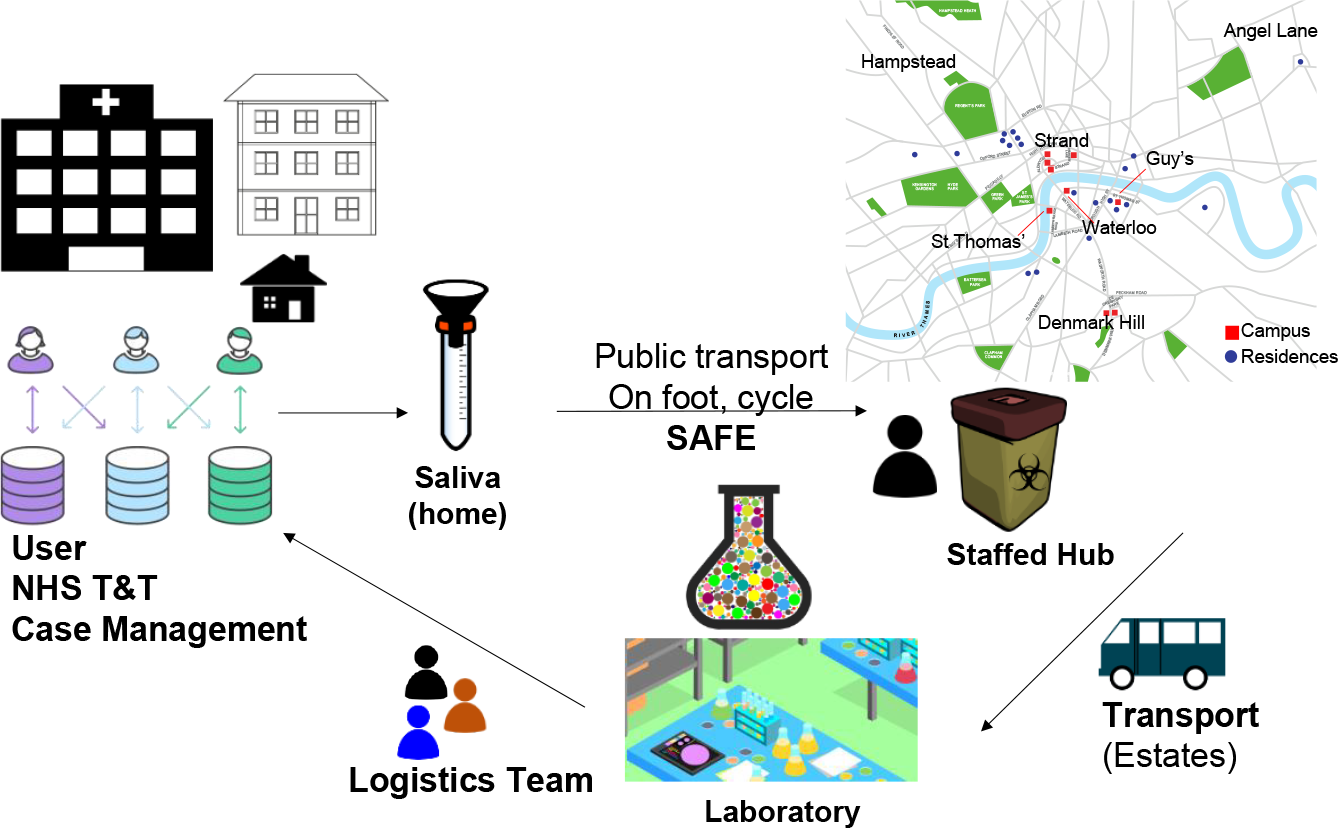
Overview of KCL TEST. Users acquired barcoded tubes in which they deposited saliva, and they brought their samples to one of our hubs (depicted in the London map). Transport of all samples occurred in 1 or 2 shifts and the laboratory processed them for SARS-CoV-2 RNA, using the same barcode as identifier. Results were sent to our online system, where they were released to the NHS Test and Trace as well as to the user and case management team to detect and prevent potential localised outbreaks.

Saliva samples were inactivated by the collection buffer as it contained 2% SDS (Supplemental Figure 1), and thus could be safely transported by users and rapidly processed by laboratory staff. In our sample kits, each saliva collection tube had a unique barcode. Users registered their samples by logging onto our online system, before dropping off the sample at a local collection point; testing was therefore unsupervised. The link between sample barcode and individual remained visible only to certain members of the case management and logistics teams.

At its peak we had18 staffed hubs in different locations across London (UK), where staff and students could collect saliva kits and drop-off their samples. Samples were collected from all the hubs and carried to the laboratory for processing. We developed our own laboratory information management system (LIMS, available as a modifiable Docker package) to log the samples using the same barcode as on the tube; this was key to reducing handling times. Samples were processed to test for the presence of SARS-CoV-2 RNA (N and E genes) and an internal human control (human *RNAseP*) using real time quantitative PCR (RT-qPCR). Laboratory members only worked with the barcode present on the tube and therefore laboratory processing remained anonymous. Results were sent to the users (matched centrally using barcode-user details) and to NHS Test and Trace. Our case management team also received information about positive individuals and risk assessed for potential outbreaks, e.g., if positive samples clustered at specific locations.

### KCL TEST Laboratory

KCL TEST evolved over time, starting with a group of 11 researchers and an operational team of 3 people (Figure 2A). KCL TEST was organised into testing, research (to develop/improve protocols) and management teams, stabilising at 10 testing team members in mid-April 2021. From then onwards, we established three working shifts that covered 24 h, 6 days per week. The night shift was on call to provide cover for possible repeats or delays. There was a 30 min overlap between shifts to improve handover efficiency between teams. These shifts grew to 17 lab members in total from October 2021 until May 2022 when we scaled down to 9 lab members and two shifts with a 3 hour overlap to cover the busiest times until July 2022.

**Figure 2.**
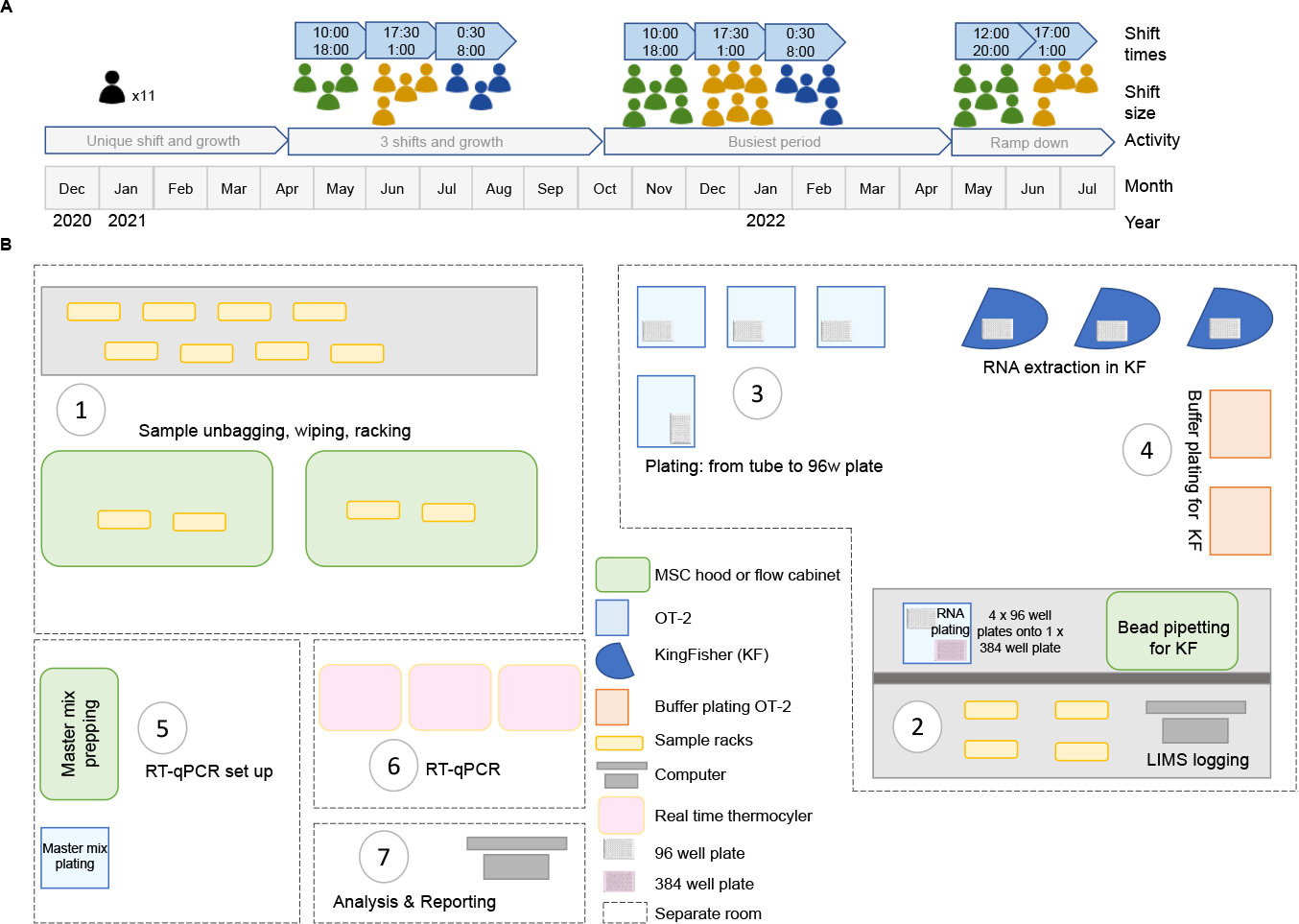
KCL TEST laboratory set up. **A**. Schematic timeline of KCL TEST laboratory, size of shift patterns and time of the shifts. **B**. Schematic diagram of our laboratory set up, with different spaces/rooms (dotted lines). Sample unbagging, wiping and racking (1) happened in one area; logging (2), plating by OT-2 robots (3) and extracting RNA (3) happened in another area, with 2 OT-2 robots plating buffers for RNA extraction with the Kingfisher Flex (KF) system. RT-qPCR was set up in a separate room to avoid contamination (5) with one master mix plating OT-2 and a sample plating OT-2 using an 8-channel multichannel pipette set out to dispense in a 384 well format. Plates were spun and taken to the thermocyclers (6) for RT-qPCR. Data were analyzed and results sent (7).

Our laboratory pipeline is summarised in Figure 2B and detailed in Supplemental Figure 2. On arrival at the laboratory samples were visually processed for leaks, unbagged, wiped and racked (1), logged in using our LIMS (2) and plated using OT-2 (Opentrons) robots onto 96 deep-well plates (3). In the meantime, buffer plating OT-2s prepared the 96 well plates required for RNA extraction (4). While RNA plates were prepared, the RT-qPCR reaction mix was calculated, prepared and aliquoted by another OT-2 onto 384 well plates. Samples from 4 x RNA plates were then plated onto 1 x 384 well plate (5) which were sealed and spun prior to the RT-qPCR run (6). Lastly, data analysis and reporting occurred (7). Every shift completed a securely shared data sheet to determine the weekly and monthly statistics (number of positives, negatives, inhibitory -also called voids- and samples).

As a critical part of our internal controls, we ensured accuracy of our OT-2 robots (Supplemental Figure 3), as well as included negative controls (PBS used in diluting saliva samples and water) and positive controls (positive internal control prepared in house) in every plate. We also ran cross-plate contamination assessments (Supplemental Figures 4 and 5) with known checkerboard and interspersed positive samples, which also enabled comparisons of the results from combining our 3 Kingfisher Flex robots with our 3 real time thermocyclers. This approach identified potential performance problems with instrumentation which we fixed, and such checks not only ensure consistent results but are also required for ISO15189 accreditation.

We initially performed RT-qPCRs in single-plex before moving to duplex detection of N gene and human *RNAseP*, finally setting in our triplex assay (viral N and E, human *RNAseP*). We used the primer sequences reported by the USA Centers for Disease Control and Prevention (CDC) ^21^. We established thresholds of detection and Ct values based on data collected from hundreds of samples to establish the optimal range of human *RNAseP* amplification. We decided to include *RNAseP* in our assay to determine with confidence that negative samples were true negatives, i.e. that the sample was positive for human saliva and negative for SARS-CoV-2 as opposed to *void* which gave no signal for human RNAseP indicating insufficient, lack of or inhibitory sampling. Our thermocycling thresholds are explained in Supplemental Table 1. Annotating these is essential, as small changes in thresholds can lead to changes in several Ct values due to the exponential nature of PCR. Laboratories should thus determine and report their thresholds of amplification so that their Ct values can be compared over time and/or between different laboratories. It was therefore imperative that the team members understood the impact of threshold differences and how important it was to consistently set out these parameters in SOPs.

We performed competency evaluations of our laboratory team that included theoretical questions about our assays as part of our accreditation requirements.

We employed a commercial standard to determine our limit of detection, which we established at 100 copies/mL by consistent amplification of both viral genes (N and E) in all 6 replicate experiments (Supplemental Figure 6). Our parameters for determining positive, negative or void are set out in Supplemental Table 2.

### Homebrew matches commercial extraction of RNA for saliva SARS-CoV-2 detection

We developed our in-house extraction method for nasal swabs called *homebrew* ^11^. We further developed this method for RNA extraction from saliva and compared its performance in 980 samples (Figure 3A and Supplemental Table 3), with homebrew slightly outperforming RNAdvance Viral XP (Beckman, BM) in sensitivity of positive cases detection as well as reduced void samples (Supplemental Table 3).

**Figure 3.**
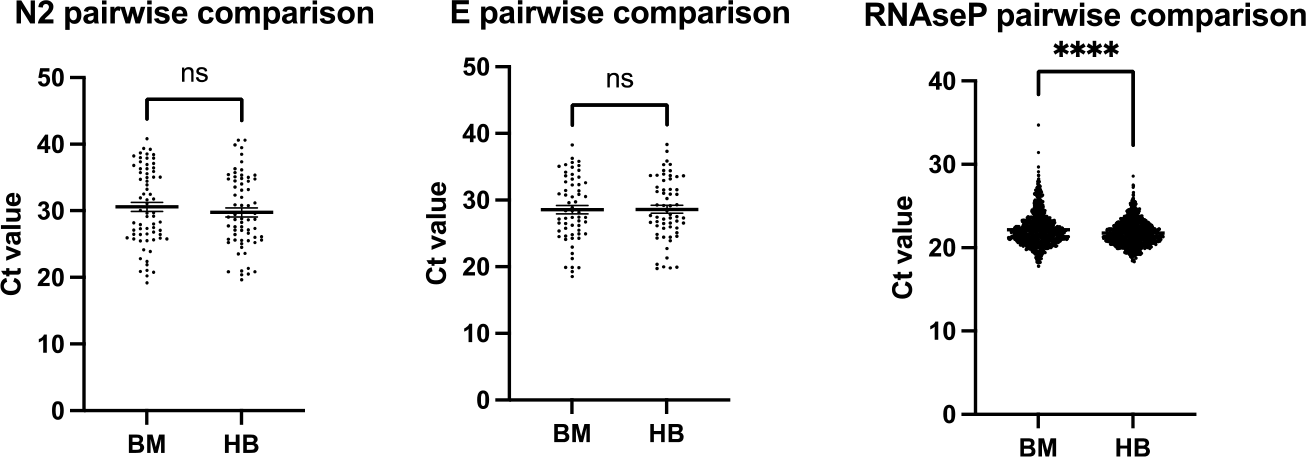
Homebrew RNA extraction matches data from commercial kits. RT-qPCR results comparing Ct values from the same samples extracted with homebrew (HB) or Beckman (BM) (n=980). **** p<0.0001 Wilcoxon two-tailed tests.

Homebrew overcame precipitation issues observed when combining sample material containing SDS and guanidinium isocyanate present in most initial lysis buffers from RNA extraction commercial kits, used to both solubilize and inactivate samples. We also developed our own positive extraction control containing inactivated SARS-CoV-2 and human cells to reflect reliably the presence of human material (*RNAseP*) as well as SARS-CoV-2 RNA. We employed molecular reagents for RT-qPCR and primers as previously ^2^. Our data (Figure 3) demonstrated that non-commercial reagents offered a comparable performance at a fraction of the price of commercial ones ^9^ and can be used and accredited for diagnostic purposes.

### Sensitivity, specificity and comparison with swab data

We also determined the sensitivity and specificity of our assay. To this end, we performed several comparisons. Firstly, we tested 150 positive and 250 negative samples employing a commercial kit (ProLab) and compared our results to that of a UKAS-accredited laboratory (St Thomas’ Hospital) (Figure 4A). 67 of these samples were also run on our in-house protocol, achieving 100% sensitivity and specificity, with both assays testing 39 positives and 28 negatives (Figure 4B).

**Figure 4.**
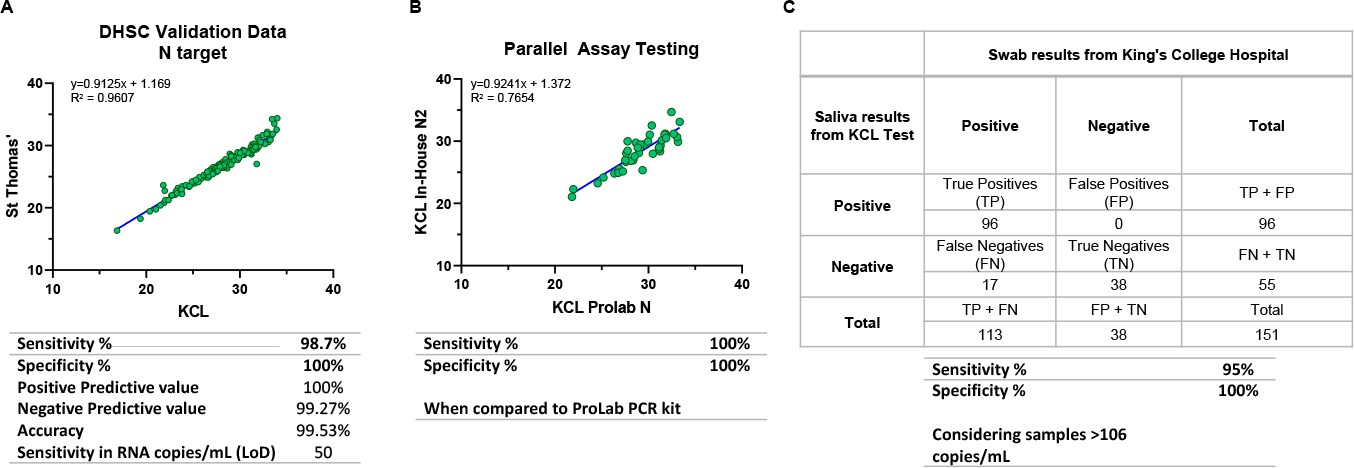
Sensitivity and specificity of KCL TEST. **A**. Linearity data on the detection of Nucleoprotein (N) viral target employing Prolab’s VIASURE SARS-COV-2 Real Time PCR Detection Kit in saliva samples. **B**. Correlation of the Ct obtained for N amplification in positive samples analysed with Prolab (Viasure) versus in-house assay. **C**. Summary of the clinical validation of our in-house assay comparing saliva vs swab.

Although saliva has been previously determined to be a valid sample for SARS-CoV-2 RNA detection, it is less used than the more established combined nose and throat swabs. We therefore performed a comparison between swab and saliva from the same individuals, with swabs being processed in another UKAS-accredited laboratory (King’s College Hospital). Our assay showed a sensitivity of 85% and specificity of 100% in accordance with the UK Government guidelines for Healthcare and public health screening and testing ^22^. Our positive count included what we called ‘inconclusive’ results as those were labelled as ‘positive at the limit of detection’ by the King’s College Hospital laboratory. When we restricted the analysis to samples with over 106 copies/mL, the sensitivity was 95% and specificity was 100%.

We assessed if some of the discrepancies that we observed with the swab data (Figure 4C) were due to different viral dynamics in swab vs saliva, defined as the duration of detectable SARS-CoV-2 RNA in the samples. To this end, we performed longitudinal sampling of a subset of positive individuals, mainly during the delta wave. Our data showed that SARS-CoV-2 detection was different between combined nose and throat swabs and saliva samples, with saliva detection showing a trend to drop faster than that in combined nose and throat swabs. (Supplemental Figure 7). This is possible due to a more rapid clearance of the virus from the different mucosae, suggesting that saliva sampling may be a more accurate way of determining infectiousness of individuals vs swabbing ^20 23^. We also recognize that this is a small sample population that mainly consisted of the delta variant; it is possible that different variants present different sample kinetics^24^.

### KCL TEST data matches with that of the UK Office for National Statistics

Over the 18-month period in which KCL TEST operated, we tested 158,277 samples. Our coverage varied between 10 and 30% of campus footfall, and 2,989 positive samples (1,89%) were reported. KCL TEST showed an increased uptake during 2021, peaking before Christmas 2021 time (Figure 5A). Testing dropped during Christmas and increased sharply again January-March 2022, when it started to drop until mid-April 2022 following a change in policy about testing. We then received fewer samples, very likely our ‘super users’. The percentage of positive samples followed a very similar pattern to that reported by the UK Office for National Statistics (ONS) (Figure 5B), except for a nearly absent delta wave, despite our testing numbers being steady and the ‘back to campus’ gatherings that occur during September. Our data also suggest that there was a sharp increase in the number of positive cases during the end of May and June 2022.

**Figure 5.**
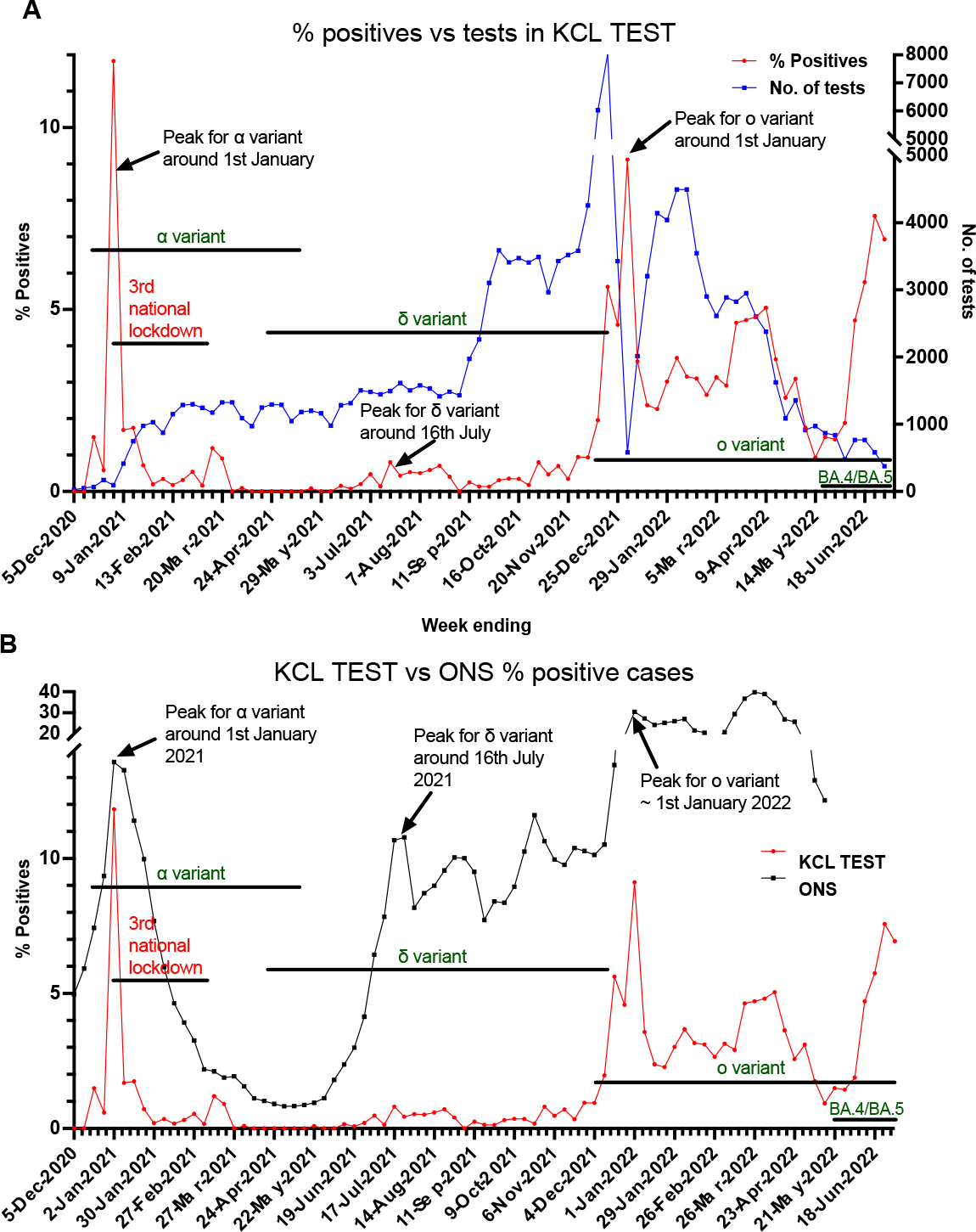
KCL TEST summary data and comparison with UK Office for National Statistics. **A**. Graph showing the longitudinal cases vs number of samples in KCL TEST. **B**. Graph showing the overlay of % positive cases detected by KCL TEST vs those reported by the UK Office for National Statistics. In both A and B we have overlayed the information on lockdowns and most common variants present at the time.

## Discussion

We present KCL TEST, our asymptomatic community surveillance programme for the detection of SARS-CoV-2 RNA. KCL TEST ran within our academic institution for over 18 months, performing 158,277 PCRs in saliva offered for free to all users. While KCL TEST ran, we developed novel protocols that offered resilience and also underwent UKAS accreditation, a requirement for test providers in the United Kingdom. We are making our molecular and automation methods available for anyone to reproduce our affordable and simple laboratory setting anywhere in the world. We are also sharing our experience and observations to promote better preparedness for the outbreaks and pandemics that will inevitably occur in the future.

‘Test, test, test’ was one of the strongest messages from the World Health Organization (WHO) in March 2020 ^25^ when COVID-19 was declared a pandemic. There was a worldwide effort to deploy laboratories, and many academic centres repurposed some of their space into diagnostics facilities^4-8 10 19^. Our and others’^26^ data call for the proactive inclusion of academic centres in pandemic preparedness. We have the facilities, skilled staff and workplace flexibility, and often have close contact with hospitals and healthcare settings. The COVID-19 pandemic revamped closer collaborations between clinical work in the hospitals and research and development in the universities, which in the past resulted in a melting pot for assay development. Maintaining these cross-disciplinary partnerships would be beneficial, especially considering the risks of future pandemics. We are keen learners and used to generating networks to improve outcomes, and cooperation between facilities is essential to expand testing capacity and share best practice ^27 28^. Our data show that we can effectively generate entire pipelines (Figures 1-3) that mirror population-wide testing (Figure 5). We should act now to generate national and international networks that can be rapidly deployed in the next pandemic.

Considering potential future pandemics and preparedness, sharing resources and expertise is crucial. A lack of reagents is avoidable and should not reoccur ^1 2^. Instead, we should cross-validate and incorporate novel pipelines that offer resilience and are proven to work ^11-15^. This is of particular importance in resource-limited settings. Health disparities based on socioeconomic status are widespread in many diseases with COVID-19 being an example. Areas of lower socio-economic indexes have the least uptake of testing but a higher proportion of positive rates and burden ^29-31^. More affordable pipelines, such as ours, can and should be available to others. This requires a more agile process for the validation and accreditation of protocols and facilities. In the UK, this is undertaken by UKAS and international bodies include International Laboratory Accreditation Cooperation (ILAC), International Accreditation Forum (IAF) or the European Accreditation (EA). Pandemic preparedness should include an emergency body that can focus on rapid establishment of diagnostic facilities to accelerate and ensure quality reporting^32^. Our internal controls and equipment-combination testing (Figure 2 and Supplemental Figures 3 and 4) highlight the importance of performing monthly checks on the different equipment combinations, together with appropriate negative and positive controls in each run, to avoid potential sources of erroneous results.

Dedicated facilities for community testing in vulnerable settings such as care homes or healthcare centres should be prioritised. However, testing should include asymptomatic individuals and be accessible to all; these objectives were at the core of KCL TEST and underpin why we chose to both employ saliva as sample source and develop low-cost pipelines. Testing requires a compromise between costs and benefits; health-wise, reducing community transmission inherently leads to less hospitalizations ^33 34^. Balancing the numbers between lost days and test costs favours the latter ^35^. Enabling affordable and reliable asymptomatic testing with adequate measures that protect workers’ health and wellbeing must be at the forefront of pandemic management. This should be part of employer’s responsibilities, i.e. consider its inclusion in Control of Substances Hazardous to Health Regulations (COSSH): indeed, a fit workforce is a productive workforce.

KCL TEST was free for anyone affiliated with King’s College London and was funded and widely publicized by our Institution, which improved uptake and contributed to adherence. We recommended testing 2 days per week for those on campus regularly, and we believe this contributed to the nearly absent delta wave (Figure 5), together with widespread immunization. We received over 12,500 Fit to Fly requests, but only 335 Day 2 and 10 Day 8 tests, suggesting that our community travelled safely and that it consisted mainly of vaccinated individuals. We also observed a drop in testing from April 2022 despite our facility maintaining capacity. We believe that these data clearly reflect the power of public messaging as free testing ended in April 2022 in the UK. This was a controversial measure^36^ and our data show that infections kept rising in the months after. However, we were not able to access weekly ONS data since the end of May 2022.

In summary, we believe our findings and set up at KCL TEST strongly endorse the inclusion of academic centres in pandemic preparedness and asymptomatic population-level testing. Our framework allows for minimally invasive sampling, rapid reporting and low-cost diagnostics. Over 20,000 KCL staff, students and household contacts benefited from our tests. Putting similar systems in place that will enable the rapid and economical mobilization of accessible testing, particularly in communities close to care homes and hospitals where avoidance of outbreaks can save lives, should be a priority, both in the UK and internationally.

## Materials and Methods

KCL TEST was initially set up as a research project under King’s College London ethics number HR-20/21-21150 and then continued as a service delivery.

Sample preparation can be briefly summarised as follows: saliva was self-collected by participants in GeneFix tubes (1mL or 2mL, Isohelix) and logged by the participants. 500μl of phosphate buffered saline (Thermofisher Scientific) were initially added to each saliva sample using OT-2 robots, and mixed to reduce viscosity. 200μl of each diluted saliva sample were plated onto deep well 96-well plates. Proteinase K (Merck) was manually mixed and incubated with each sample to digest mucins and further reduce viscosity. RNA binding SpeedBead Magnetic Carboxylate Modified Particles (Cytiva) were manually added as a solution containing NaCl and Isopropanol. Kingfisher Flex robots (Thermofisher Scientific) were then used to isolate RNA. One-step quantitative real-time PCR was performed using Luna® Probe One-Step RT-qPCR 4X Mix with UDG (New England Biolab). The primer/probe sequences from the US CDC for SARS CoV-2 N2 and *RNaseP* genes, and Charité PHE E, Sarbeco for the SARS CoV-2 E gene were used (Integrated DNA Technologies).

Full details of methods and protocols are provided in SOP001 and SOP002. Opentrons OT-2 liquid handling robot python scripts, a blank SOP (for reference) and competency assessments for laboratory staff are also provided. We have made all our Standard Operating Procedures, scripts and LIMS available under a CC BY NC license in Open Science Framework upon request. The LIMS should be tailored to each individual set up (e.g. considering the number of robots dedicated to each task, 96 vs 384 well plate layout).

This work is available under a CC-BY-NC license.

## Competing Interests

Dr Rahul Batra is the owner of IC Intelligence LTD who helped with the processing of samples for UKAS accreditation.

## Author Contributions

J.R.A., E.S., S.P-S., K.P., T.A.R, P.K., G.D., S.O., I.I., N.G., I.G., E.C., T.J.A.M. contributed with acquisition, analysis and interpretation of the data. A.M.O-P and J.J.G., A.V.F.T., A.G. and S.W. contributed with acquisition and analysis of data. R.B. contributed with data acquisition. S.P., A.M-V. contributed with data analysis. C.T. and M.K. created essential infrastructure for KCL TEST. D.L., L.T., A.G., and V.G. contributed by creating new software and scripts used in our work. S.N, R.P.R.G. and H.M. provided supervision. J.M., C.C., M.Z., R.R., M.H.M. and R.T.M-N conceived, designed and supervised the work. All authors approved the manuscript.

## Funding and Acknowledgements

We are incredibly grateful to the whole King’s College London community for their support and uptake of KCL TEST. We are also very grateful to Prof Chris Denning, Prof Jonathan Ball and Prof Moira Petrie at the University of Nottingham for sharing their experience and their advice on UKAS accreditation. We are very thankful to David Sherrin and his team for helping us with all speedy laboratory refurbishments. KCL TEST was funded by King’s College London and supported by the Huo Family Foundation grant (MHM and RTMN).

## Notes

### Funding Statement

KCL TEST was funded by Kings College London and supported by the Huo Family Foundation grant (MHM and RTMN).

### Author Declarations

KCL TEST was initially set up as a research project under Kings College London ethics number HR-20/21-21150 and then continued as a service delivery.

### Summary of Updates

New Abstract, addition of competing interests and author contributions

