## Supplemental Figures and Tables for "KCL TEST: an open-source inspired asymptomatic SARS-CoV-2 surveillance programme in an academic institution"

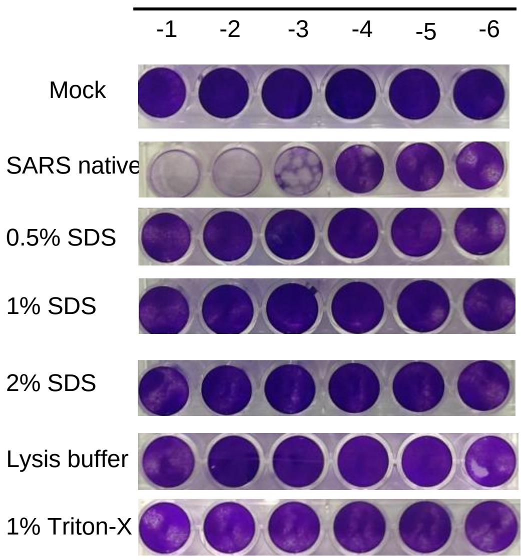

**Supplemental Figure 1. Plaque assays demonstrating inactivation of SARS-CoV-2 in saliva**. Our collection tubes contained 2% SDS and thus we tested decreasing concentrations of the detergent on plaque assays, using increasing amounts of SDS, buffer or 1% Triton-X. Vero E6 TMPRSS2 were employed. All samples spun through size exclusion columns (Amicon) 3 times to remove any detergent contaminants that would also kill cells.

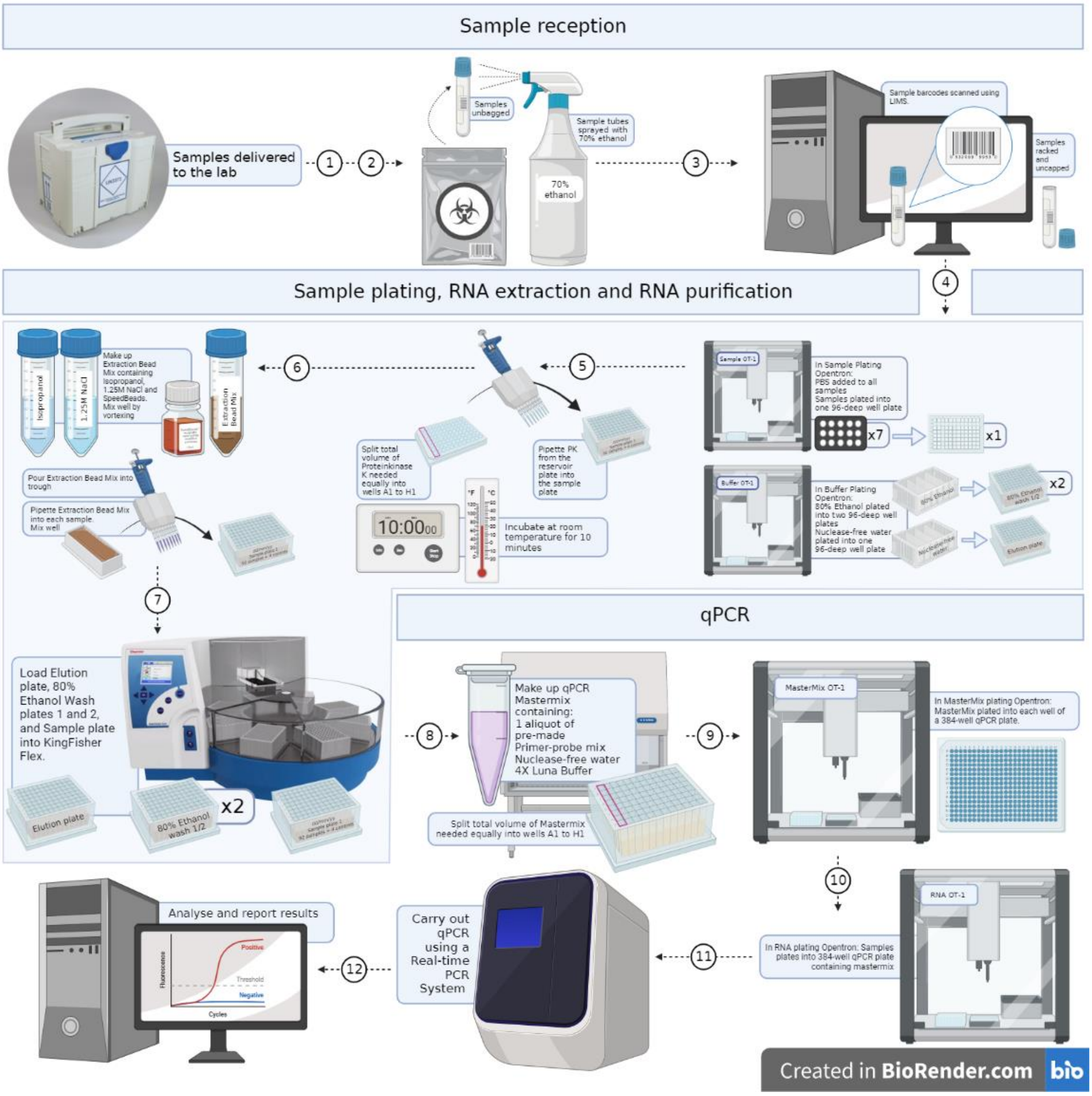

**Supplemental Figure 2. Detailed diagram of our testing pipeline.** Detailed step by step diagram of our workflow in the laboratory, from sample reception to result reporting.

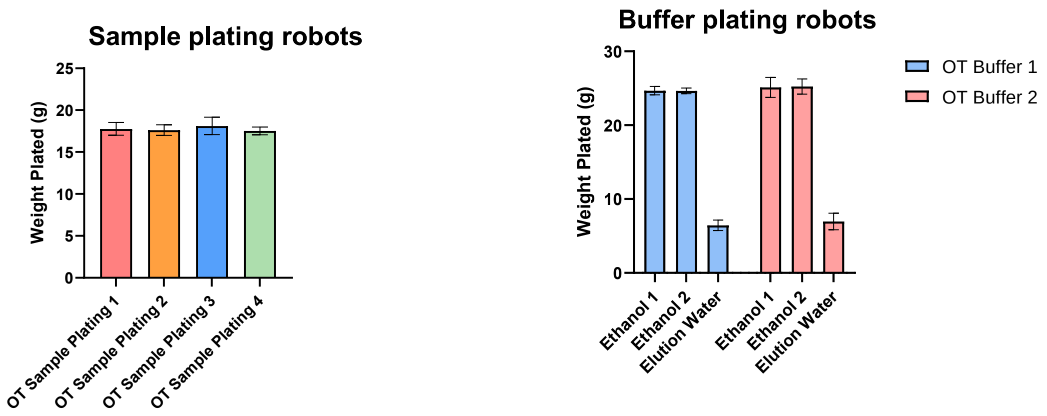

**Supplemental Figure 3. Accuracy of sample plating and buffer plating OT-2 robots.** Each OT-2 was tested over 3 days and filled plates were weighted to determine if different machines had different accuracy. Our data showed that all robots performed equally in their plating.

| | 1 | 2 | 3 | 4 | 5 | 6 | 7 | 8 | 9 | 10 | 11 | 12 |
|---|---|---|---|---|---|---|---|---|---|---|---|---|
| A | **H2O** | Neg | Neg | Neg | Neg | Neg | Neg | Neg | Neg | Neg | Neg | Neg |
| B | Neg | Neg | Neg | Neg | Neg | POS 1 | Neg | Neg | POS 10 | Neg | Neg | Neg |
| C | Neg | POS 4 | Neg | Neg | Neg | Neg | Neg | Neg | Neg | Neg | Neg | Neg |
| D | Neg | Neg | Neg | POS 7 | Neg | Neg | POS 8 | Neg | Neg | Neg | POS 3 | Neg |
| E | Neg | Neg | Neg | Neg | Neg | Neg | Neg | Neg | POS 6 | Neg | Neg | Neg |
| F | Neg | POS 2 | Neg | Neg | Neg | POS 5 | Neg | Neg | Neg | Neg | Neg | Neg |
| G | Neg | Neg | Neg | Neg | Neg | Neg | Neg | POS 9 | Neg | Neg | Neg | **$H_20$** |
| H | Neg | Neg | Neg | Neg | Neg | **$H_20$** | Neg | Neg | Neg | Neg | Neg | **PEC** |

**Supplemental Figure 4. Example Interspersed Positive Plate and respective colour key.** These layouts were repeated for all combinations of our Kingfisher Flex RNA extractors and thermocyclers. Neg: negative sample, POS positive sample, PEC Positive Extraction Control.

| | | | | | | | | | | | |
|---|---|---|---|---|---|---|---|---|---|---|---|
| - | 25.58 | - | 28.30 | - | 25.19 | - | 23.90 | - | 26.39 | - | 23.97 |
| 24.85 | - | 26.02 | - | 25.28 | - | 24.46 | - | 23.82 | - | 25.36 | - |
| - | 25.55 | - | 25.75 | - | 25.33 | - | 24.07 | - | 24.91 | - | 23.85 |
| 26.12 | - | 25.39 | - | 26.51 | - | 24.10 | - | 24.23 | - | 23.85 | - |
| - | 25.81 | - | 25.35 | - | 25.29 | - | 23.84 | - | 24.00 | - | 25.15 |
| 24.95 | - | 25.84 | - | 25.58 | - | 23.90 | - | 25.84 | - | 24.82 | - |
| - | 25.34 | - | 24.21 | - | 24.24 | - | 24.25 | - | 24.31 | - | - |
| 25.10 | - | 25.45 | - | 25.26 | - | 25.49 | - | 23.83 | - | 23.97 | 31.52 |

**Supplemental Figure 5. Example Checkerboard Experiment for adjacent contamination, target Ct 26.** These layouts were repeated for all combinations of our Kingfisher Flex RNA extractors and thermocyclers.

Limit of Detection with SARS-CoV-2 Analytical Q Panel 01 (SCV2AQP01-A)

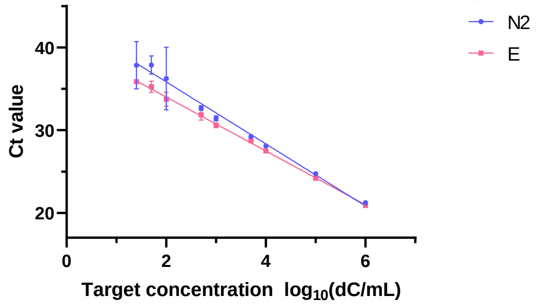

| | Equation |
|---|---|
| N2 | Y = -3.751*X + 43.34 |
| E | Y = -3.257*X + 40.52 |

| | R squared |
|---|---|
| N2 | 0.9267 |
| E | 0.9913 |

| | | | | N2-SUN 3plex MM | | | | |
|---|---|---|---|---|---|---|---|---|
| | | | | N2_SUN | | E_FAM | | |
| Sample number | ID | Concentration | Log10 Concentration (dC/mL) | Mean Ct | SD | Mean Ct | SD | Result |
| 1 | SCV2AQP01-S01 | 1,000,000.00 | 6.00 | 21.24 | 0.14 | 20.91 | 0.15 | Positive |
| 2 | SCV2AQP01-S02 | 100,000.00 | 5.00 | 24.73 | 0.22 | 24.20 | 0.14 | Positive |
| 3 | SCV2AQP01-S03 | 10,000.00 | 4.00 | 28.09 | 0.12 | 27.48 | 0.09 | Positive |
| 4 | SCV2AQP01-S04 | 5,000.00 | 3.70 | 29.25 | 0.13 | 28.74 | 0.22 | Positive |
| 5 | SCV2AQP01-S05 | 1,000.00 | 3.00 | 31.44 | 0.32 | 30.58 | 0.29 | Positive |
| 6 | SCV2AQP01-S06 | 500.00 | 2.70 | 32.66 | 0.28 | 31.84 | 0.65 | Positive |
| 7 | SCV2AQP01-S07 | 100.00 | 2.00 | 36.25 | 3.78 | 33.73 | 0.85 | Positive |
| 8 | SCV2AQP01-S08 | 50.00 | 1.70 | 37.88 | 1.09 | 35.23 | 0.67 | Positive |
| 9 | Qnostics '10' | 25.00 | 1.40 | 37.84 | 2.85 | 35.86 | 0.11 | Positive |
| 10 | SCV2AQP01-S09 | 0 (Negative) | - | - | - | - | - | Negative |

**Supplemental Figure 6. Limit of detection in KCL TEST employing a commercial standard.** The Qnostics panel used (SARS-CoV-2 Analytical Q Panel 01: SCV2AQP01-A) is a viral suspension which was extracted and amplified as per our sample pipeline. The standards provided range in concentrations between 1 million and 50 digital copies/mL. The lowest standard available was diluted further to provide a sample with approximately 25 copies/mL. Extractions and RT-qPCRs were repeated n=6 times. All 6 replicates showed amplification of at least one of the targets (N2 or E) for all concentrations up to 25 copies/mL. At the lowest concentration, 25 copies/mL, amplification for both targets (N2 and E) were only detected in 1 out of 6 replicates and at 50 copies/mL on 3 out of 6 replicates. There were more replicates with amplification of N2 compared to E. The mean Ct for each concentration can be seen in the table below. The mean Ct values for E and N2 between 25 and 50 copies/mL were 35 and 38, respectively.

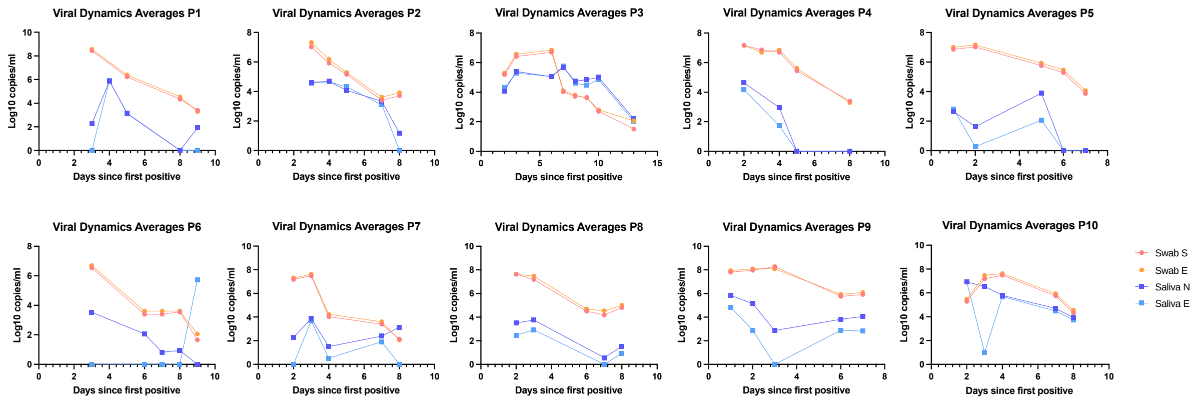

**Supplemental Figure 7. Viral dynamics comparing combined nose and throat swab vs saliva**. We observed a general trend towards having less copies per mL in saliva as compared with swabs.

### Supplemental Tables

| Target Thresholds | |
|---|---|
| N2 | 0.02 |
| E | 0.02 |
| RNAseP | 0.02 |

**Supplemental Table 1. Thresholds employed in In-house Triplex SARS-COV-2 Real Time PCR**. Thresholds set to determine Ct values after RT-qPCR.

| TARGET | Positive | Positive at LoD | Negative | Inhibitory /Void | Inconclusive |
|---|---|---|---|---|---|
| N2 and E | <36 for both targets | >36 for both targets | Undetermined | | |
| N2 or E | <36 for one target | | | | ≥36 for one target |
| RNase P | <26.5 | <26.5 | <26.5 | ≥26.5 | <26.5 |

**Supplemental Table 2. Key to interpret our RT-qPCR results.**

| | Beckman Kit | Homebrew |
|---|---|---|
| **Number of values** | 980 | 980 |
| **Positive** | 61 | 66 |
| **Negative** | 907 | 906 |
| **Inconclusive** | 8 | 7 |
| **Void** | 4 | 1 |

**Supplemental Table 3. Comparison of homebrew vs Beckman extraction**. Sample characteristics and number of detected positives/negatives and voids.
